# Neutrophils cultured *ex vivo* from CD34^+^ stem cells are immature and genetically tractable

**DOI:** 10.1101/2023.07.12.23292345

**Authors:** Claire A. Naveh, Kiran Roberts, Christopher M. Rice, Kathryn Fleming, Megan Thompson, Nawamin Panyapiean, Stephanie Diezmann, Pedro L. Moura, Ashley M. Toye, Borko Amulic

**Affiliations:** School of Cellular and Molecular Medicine, Biomedical Sciences Building, University of Bristol, Bristol, BS8 1TD, UK; School of Biochemistry, Biomedical Sciences Building, University of Bristol, Bristol, BS8 1TD, UK; Center for Hematology and Regenerative Medicine, Department of Medicine Huddinge (MedH), Karolinska Institutet, Huddinge, Sweden

**Keywords:** Neutrophils, development, haematopoietic stem cells, *ex vivo* culture

## Abstract

Neutrophils are essential antimicrobial effector cells with short lifespans. During infection or sterile inflammation, accelerated production and release of immature neutrophils from the bone marrow serves to boost circulating neutrophil counts. To facilitate the study of neutrophil development and function, we optimised a method for *ex vivo* production of human neutrophils from CD34^+^ haematopoietic progenitors. We obtain high yields of neutrophils, which phenotypically resemble immature neutrophils released into the circulation upon administration of GCSF to healthy donors. We show that *ex vivo* differentiated immature neutrophils have similar rates of ROS production but altered degranulation, cytokine release and antifungal activity compared to mature neutrophils isolated from peripheral blood. We demonstrate that *ex vivo* cultured neutrophils are genetically tractable via genome editing of precursors and thus provide a powerful model system for investigating the properties and behaviour of immature neutrophils.

## Introduction

Neutrophils are the most abundant and potent antimicrobial effector cells in humans. They combat bacterial and fungal pathogens using an arsenal of antimicrobial responses, including phagocytosis, reactive oxygen species (ROS) production, degranulation and release of extracellular chromatin decorated with antimicrobial peptides, termed neutrophil extracellular traps (NETs) [1]. Patients with congenital neutropenia, who have abnormally low circulating neutrophils counts, regularly succumb to severe bacterial and fungal infections [2, 3]. In contrast, excessive or dysregulated neutrophil activity promotes pathology in sepsis [4] and malaria [5, 6], as well as in non-infectious diseases such as cancer [7], autoimmunity [8] and cardiovascular disease [9]. Regulation of neutrophil development and function is therefore essential for health.

Neutrophils develop in the bone marrow from CD34-expressing (CD34^+^) granulocyte-monocyte progenitors (GMPs) [10]. This process, termed granulopoiesis, produces an estimated 10^11^ neutrophils daily in healthy individuals [11]. Development from the committed proliferative myeloblast to release of mature neutrophils into circulation takes approximately 14 days [12], during which progenitors gradually acquire cytoplasmic granules, lobulated nuclei and ROS producing enzymes such as NADPH oxidase (NOX2) and myeloperoxidase (MPO). Granulopoiesis is driven by the growth factors granulocyte colony stimulating factor (GCSF) [13, 14] and granulocyte-macrophage colony-stimulating factor (GM-CSF) [15, 16]. In addition to promoting differentiation in the bone marrow, GCSF promotes mobilisation of mature and immature neutrophils out of the bone marrow and into the circulation, where it also extends their lifespan [17, 18].

Neutrophils were traditionally considered to be homogenous cells. This view has recently been challenged, with reports of various activation and differentiation states, both in healthy individuals [19] and during inflammation [7, 11]. One of the main determinants of neutrophil phenotype is maturity. During inflammation, elevated GCSF production promotes the release of immature neutrophils from the bone marrow. This phenomenon has been observed for decades in clinical settings, where the presence of immature morphological features in circulating neutrophils is termed ‘left shift’. Flow cytometry studies of patient blood can identify immature neutrophils by reduced expression of surface maturity markers such as CD10 and CD101, relative to mature neutrophils [20]. Strikingly, the number of circulating immature neutrophils is often associated with poor prognosis in a variety of autoimmune and inflammatory diseases such as systemic lupus erythematosus (SLE), COVID-19 and late-stage cancer [20–26]. Despite this strong association with severe disease, the function and inflammatory potential of immature neutrophils remains unclear.

GCSF administration is sufficient to mobilise immature neutrophils into the circulation; this was demonstrated in GCSF-treated allogeneic stem cell donors (GCSF-D) [27]. Importantly, these studies suggested functional differences between control and GCSF-D neutrophils, including elevated production of proinflammatory cytokines, reduced motility and capacity to produce ROS, as well as reduced ability to suppress the fungal pathogen *Candida albicans (C. albicans)* [18, 27–29]. These functional differences define a new appreciation for neutrophil heterogeneity or ‘neutrophil states’, breaking the mould of the terminally differentiated, uniform cells, which neutrophils have long been described as.

Despite their suggested role in a variety of diseases, immature neutrophils remain poorly understood. In fact, almost all biochemical and functional analyses of neutrophils performed over the last hundred years have been done on steady state neutrophils isolated from healthy donors. This knowledge gap exists because we do not have a reliable source of immature neutrophils, due to their absence in healthy donors and difficulties in accessing patient and GCSF-D samples. To address this gap, we optimised an *ex vivo* protocol for differentiation of neutrophils from human hematopoietic stem and progenitor cells (HSPCs). Previous culture protocols have described neutrophil differentiation from diverse sources such as embryonic stem cells, induced pluripotent stem cells as well as bone marrow and peripheral blood HSPCs; these studies report that cultured neutrophils resemble native neutrophils in nuclear morphology, surface marker expression and some neutrophil effector functions such as ROS production, bacterial killing, phagocytosis and chemotaxis [30–40]. Based on these reports, we optimised the culture conditions to obtain a high yield of CD34^+^ HSPC derived neutrophils. We show that cultured neutrophils more closely resemble immature neutrophils than steady state ones, and that they are amenable to genome editing, enabling mechanistic studies of immature neutrophil function.

## Results

### Optimisation of a neutrophil culture and differentiation protocol from CD34^+^ HSPCs

We isolated CD34^+^ HSPCs from peripheral blood using immunomagnetic selection and cultured these with multiple combinations of stem cell proliferation and expansion factors (stem cell factor (SCF), interleukin-3 (IL-3) and fms-like tyrosine kinase 3 ligand (Flt3-L)), followed by neutrophil differentiation factors (GM-CSF and G-CSF). Our optimised differentiation protocol, shown in Supplemental Fig 1A, was selected based on superior yield and differentiation efficiency. This protocol resulted in 326 ± 248 fold expansion (n= 7-11, Fig. S1B) with progressively increasing expression of granulocyte markers (CD66b and CD11b) and a reduction in CD34 expression (Fig. S1C). Neutrophil nuclear lobulation and surface markers peaked at differentiation day 17 (Fig. S1A and C), yielding an average of 75.45% neutrophils expressing both CD66b and CD15 (n=4, Fig. S1D).

### Cultured neutrophils phenotypically resemble GCSF-mobilised immature neutrophils

GCSF treated healthy donors (GCSF-D) are known to have a significant population of immature neutrophils circulating in peripheral blood [41]. We used flow cytometry to compare profiles of cultured, GCSF-D and steady state native neutrophils. GCSF-D and steady state native neutrophils had similar forward (FSC) and side scatter (SSC) (Fig. 1A-C), indicating similar size and granularity. Cultured neutrophils displayed elevated FSC and decreased SSC indicating decreased granularity, possibly due to reduced abundance of cytoplasmic vesicles. As expected, GCSF-D neutrophils had significantly lower expression of maturity markers CD10 and CD101 compared to native neutrophils (Fig.1D-F, Fig. S1E-F). Cultured neutrophils also had very low abundance of maturity markers, indicating immature status. In contrast, we found similar levels of granulocyte markers in all three cell types, with a trend for increased CD66b in cultured neutrophils (Fig 1. G-I). In summary, based on their surface marker expression, neutrophils cultured from CD34^+^ HSPCs phenocopy immature GSCF-D neutrophils.

**Fig. 1.**
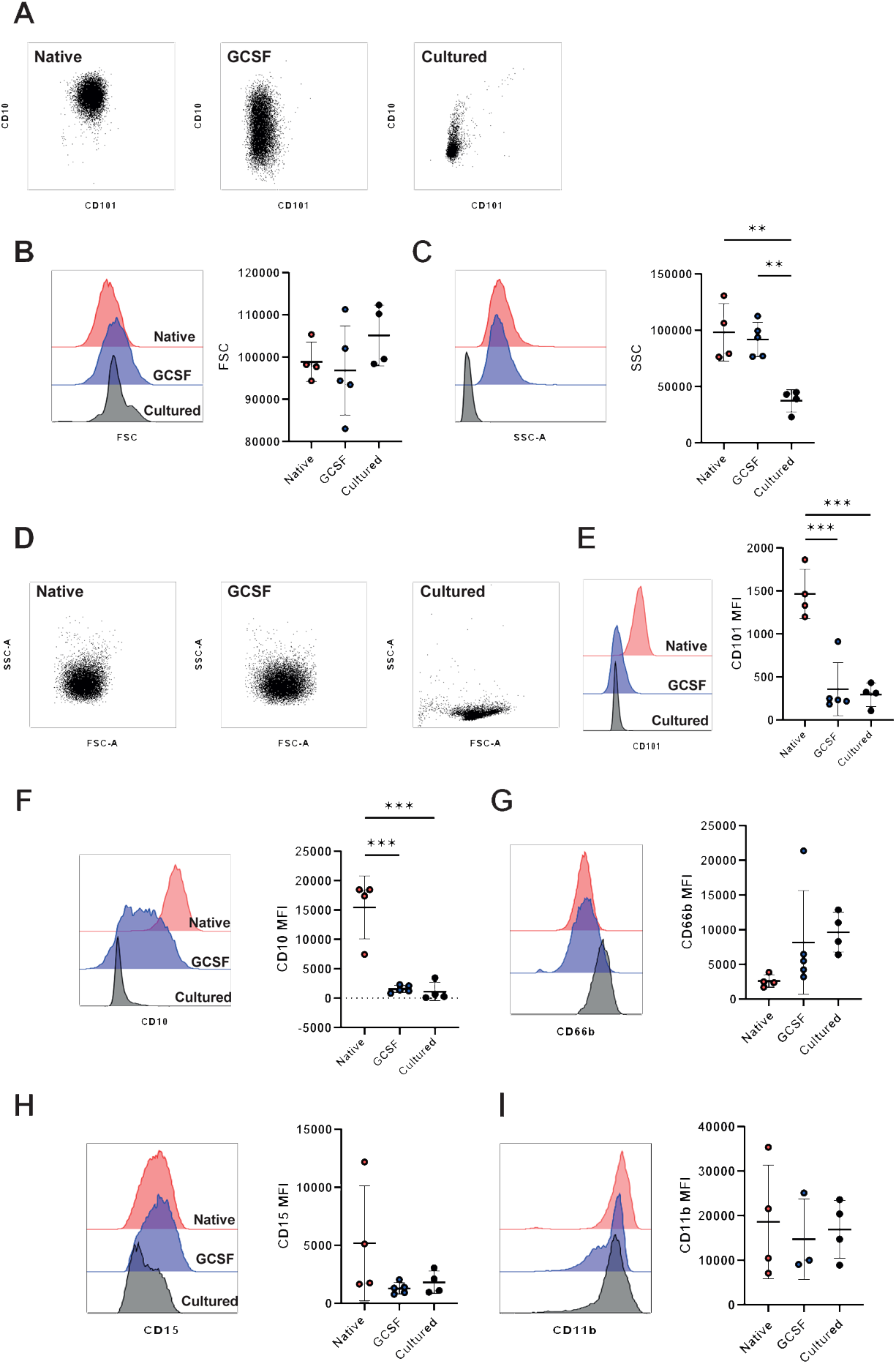
Flow cytometry analysis of surface markers on cultured, GCSF-D and control neutrophils. A) Representative scatter dot plots displaying FSC and SSC of neutrophils isolated from peripheral blood of native and GCSF treated donors (GCSF-D) or cultured neutrophils. B-C) Representative histogram and quantification of FSC (B); and SSC (C). D) Representative scatter dot plots displaying representative CD101 and CD10 expression on native, GCSF-D and cultured neutrophils. E-F) Representative histograms and mean fluorescent intensity (MFI) quantifications of CD10 (E) and CD101 (F). Histograms colour coded as: native (red), GCSF-D (blue) and cultured (black) neutrophils. G-I) Representative histogram and quantification of MFI for CD66b (G), CD15 (H), CD11b (I). Data were analysed by one-way ANOVA with Tukey’s multiple comparisons displayed on graph, n=3-5, * = p<0.05, ** = p< 0.01, *** = p< 0.001.

### Functional comparison of cultured and native neutrophils

Immature peripheral blood neutrophils are reported to have reduced oxidative burst, impaired capacity to kill *C. albicans* and increased cytokine production to toll-like receptor (TLR) agonists [27–29]. We compared effector responses in cultured neutrophils and steady state native neutrophils isolated from peripheral blood, to explore any possible functional differences. We observed no difference in total ROS production, in response to phorbol myristate acetate (PMA), a protein kinase C (PKC) agonist, when analysed by area under the curve (AUC) (Fig. 2A). The kinetic curve did however reveal subtle differences in the temporal response across donors, potentially indicating differences in in antioxidant response or NOX2 assembly. ROS detection with aminophenyl fluorescein (APF), a dye that detects peroxynitrites and MPO-catalysed hypochlorous acid, also showed comparable production of intracellular ROS (Fig. 2B). In summary, NOX2 and MPO activity are similar in cultured and native neutrophils.

**Fig. 2.**
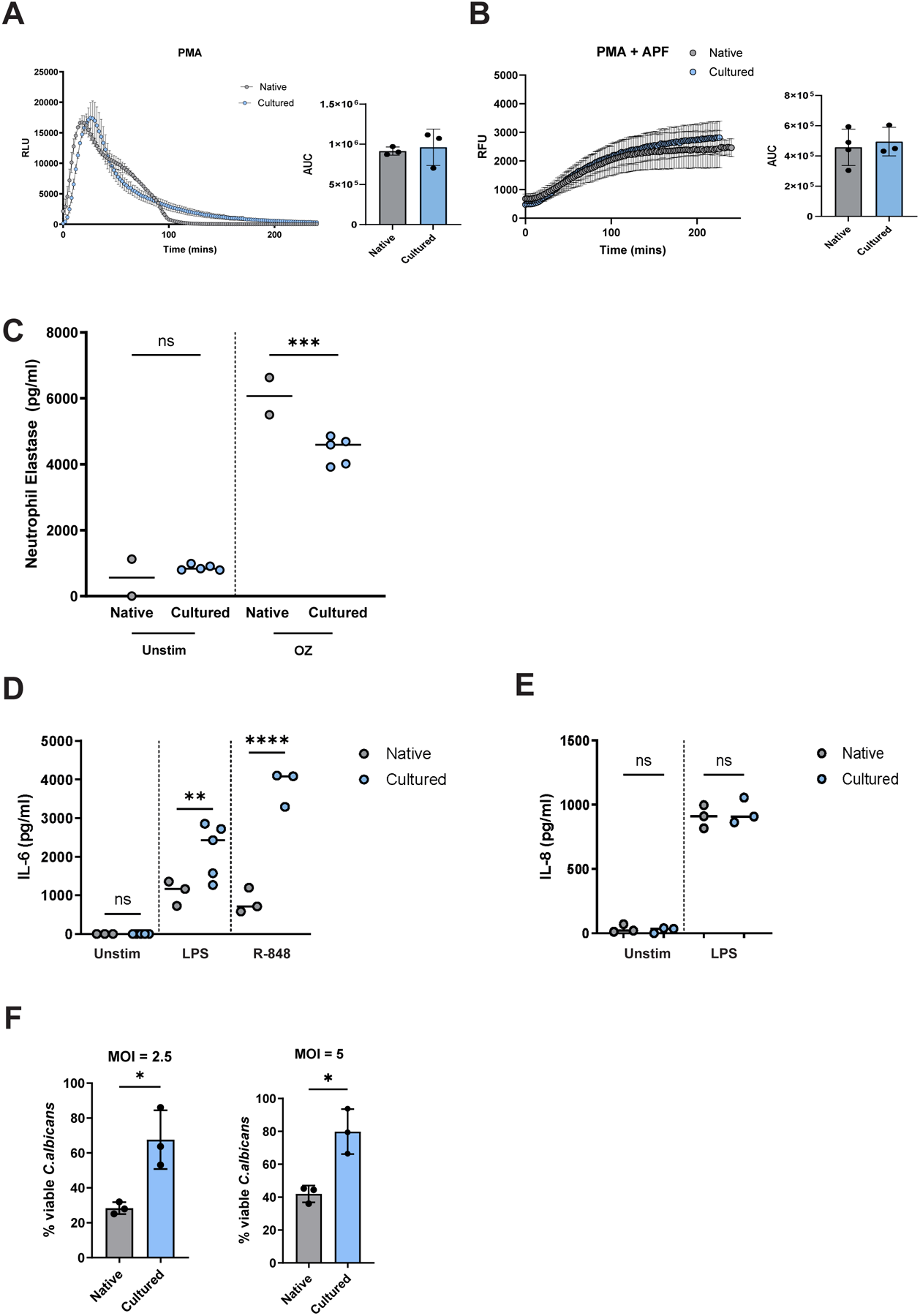
Comparison of effector functions in cultured and native neutrophils. Detection of ROS with A) luminol and B) APF in native or cultured neutrophils stimulated with 100nM PMA. Left: A representative kinetic plot of the respiratory burst in a single control donor and single differentiation; right: area under the curve (AUC) quantification. C) Exocytosis of NE in response to stimulation with 25 µg/ml opsonised zymosan (OZ) for 1 hour, quantified by ELISA. D) IL-6 and E) IL-8 cytokine release from native and cultured neutrophils stimulated overnight with 100ng/mL LPS or 5µM R-848, quantified by ELISA. F) *C. albicans* viability determined using AlamarBlue in native and cultured neutrophils incubated with opsonised *C. albicans* for 2.5h at MOI 2.5 (left) or 5 (right). Error bars indicate mean ± standard deviation, n=2-5. Data were analysed by two tailed students t test, * = p<0.05, ** = p< 0.01, *** = p< 0.001.

Next, we measured exocytosis of primary granules by quantifying neutrophil elastase (NE) release in response to stimulation with opsonized zymosan (OZ). We observed a 27% reduction in extracellular NE release in stimulated cultured neutrophils compared to native ones (Fig. 2C), suggesting either reduced NE expression or decreased propensity to degranulate in response to OZ.

As reported for immature neutrophils from GCSF-D, production of proinflammatory cytokine interleukin-6 (IL-6) was significantly elevated in cultured neutrophils in response to both TLR4 agonist lipopolysaccharide (LPS) and TLR7/8 agonist resiquimod (R-848) (Fig. 2D). However, no difference in IL-8 production was detected in response to LPS (Fig. 2E).

Lastly, we found that cultured neutrophils were able to kill the fungal pathogen *C. albicans,* however this was reduced compared to native neutrophils (approx. 50% reduction) (Fig. 2F).

### Cultured and native neutrophils have distinct proteomes

To investigate what underpins the functional differences described above, we analysed the proteomes of cultured and steady state native neutrophils, using Tandem Mass Tag (TMT) semi-quantitative mass spectrometry. Cultured and native neutrophils were obtained from the same donor, allowing for donor-matched proteomic analysis. Prior to mass spectrometry, both native and cultured neutrophils were FACS sorted for CD66b^+^ to eliminate contamination with precursors or other cell types (Fig. 3A). We applied a stringent false discovery rate (FDR) filtering step of ≤ 1%, which led to identification of 2359 proteins. 74% of detected proteins (1745 in total) were unchanged between cultured and native neutrophils (Fig. 3B). Enriched (red) and underrepresented (blue) proteins were defined by an absolute log2 fold change (Log2FC) of at least 1 and comparison p-value <0.05. Enriched and under-represented proteins were altogether consistent among all donors as visualised by heatmap (Fig. 3B) and volcano plot (Fig. 3C) of all differentially expressed proteins. Despite the change in relative abundances of proteins, only 12 proteins were exclusively detected in cultured neutrophils and not in native cells (Supplementary Table 1).

**Fig. 3.**
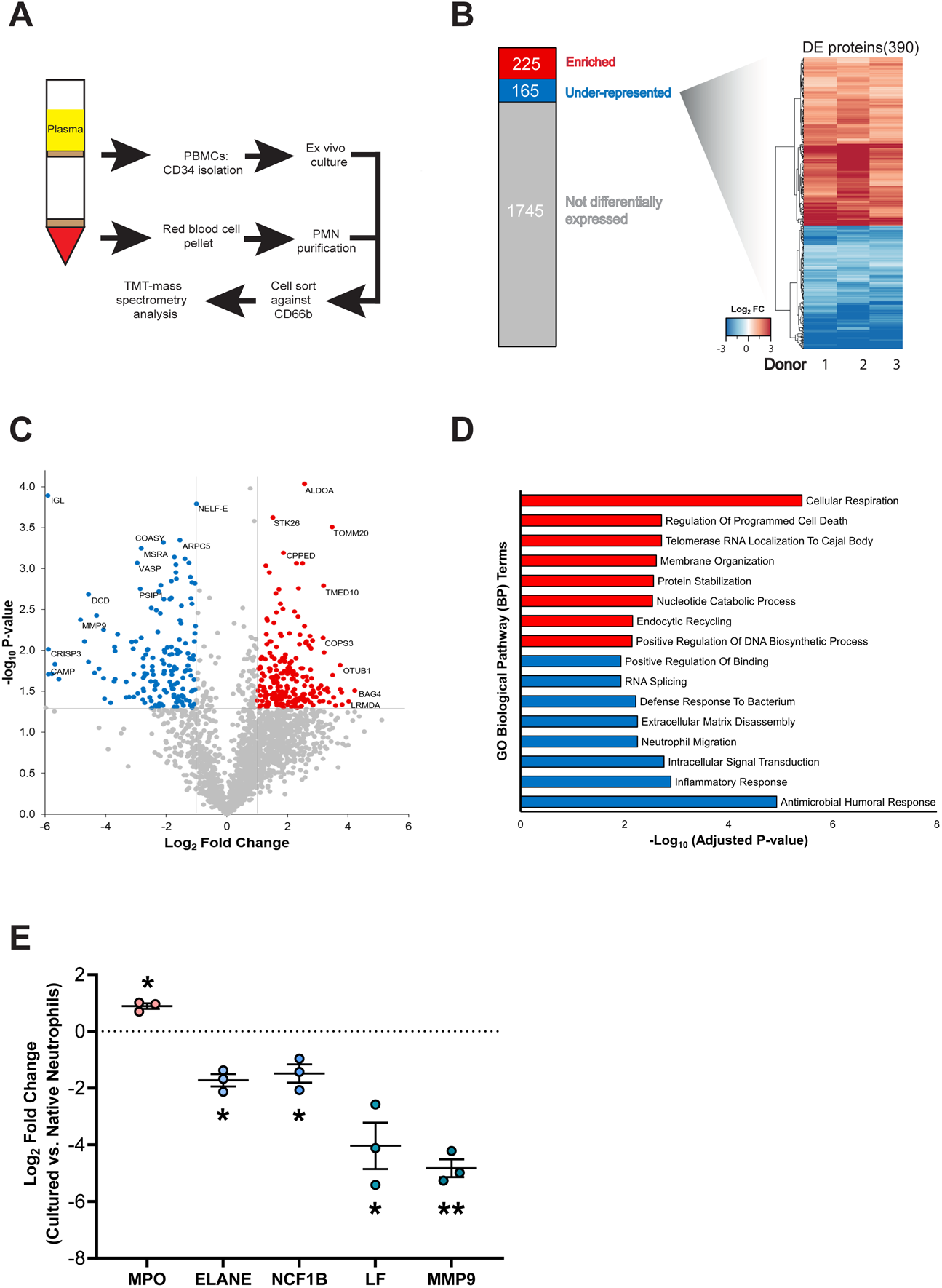
Mass spectrometry comparison of proteomes of cultured and native neutrophils. A) Experimental design for tandem mass tagged (TMT) proteomic analysis comparing cultured versus native neutrophils. B) Stacked bar chart displaying total and differentially expressed proteins between neutrophil populations, using native neutrophils as the baseline for comparison. A total of 2147 proteins were quantified by TMT, of which 225 were significantly enriched (red), 165 were significantly under-represented (blue) and 1745 were not differentially expressed between (grey) in cultured neutrophils. Differentially expressed (DE) proteins were displayed using heatmap visualisation of Log2 Fold Change (Log2FC) values of both significantly overexpressed and under-expressed proteins. C) Volcano plot displaying total and differentially expressed proteins (P-value threshold of 0.05, absolute Log2FC threshold of 1.00). D) Top 8 unique Gene Ontology (GO) terms resulting from gene list enrichment analysis of enriched (red) and underrepresented (blue) genes using the GO Biological Pathway module. E) Normalised abundances of key granule proteins, n=3 differentiations, * = p<0.05, **=p<0.001.

To identify differentially regulated pathways, gene ontology (GO) term analysis was conducted on enriched and underrepresented protein sets (Figure 3D) with the entire human proteome set as the background. This identified ‘cellular respiration’ as the top enriched pathway in cultured cells, indicating an altered metabolic state. Similarly, both ‘protein stabilisation’ and ‘nucleotide catabolic process’ were among the top 6 enriched pathways, further suggesting that biosynthetic pathways are altered in cultured neutrophils, a finding consistent with the fact that enhanced biosynthesis and mitochondrial respiration are both more prominent in neutrophil precursors, compared to mature neutrophils [42]. On the other hand, multiple antimicrobial pathways were downregulated in cultured neutrophils, including ‘antimicrobial humoral response’, ‘inflammatory response’ and ‘defence response to bacterium’, potentially explaining the impaired fungal killing.

To further interrogate specific molecular pathways altered between cultured and native neutrophils, we also carried out Reactome pathway analysis. As with the GO analysis, the top enriched Reactome pathway in native neutrophils was ‘TCA Cycle and respiratory electron transport’ (Fig. S2A), again identifying an enrichment of mitochondrial proteins in immature cultured neutrophils. STRING analysis of the 225 overexpressed proteins also highlighted a cluster of diverse mitochondrial proteins (highlighted in blue, Fig. S2B). Mitochondrial respiration is a hallmark of neutrophil precursors, as well as of immature neutrophils circulating in inflammatory disease such as COVID-19 and cancer [20, 43].

Several granule proteins stood out as significantly reduced in cultured neutrophils, including MMP9, CRISP3, CAMP and NE (Fig. 3C and Fig. 3E), supporting our findings on reduced SSC and reduced degranulation in cultured versus native cells. Reactome analysis also identified the ‘innate immune response’ and ‘neutrophil degranulation’ as the most underrepresented pathways in cultured neutrophils (Fig. S2A). Neutrophil degranulation was enriched in both overrepresented and underrepresented proteins, albeit more strongly in the underrepresented protein set (Fig. S2 A, B and C), so we further investigated granule protein abundance. Indeed, comparison of the individual abundance of representative granule proteins highlighted significant reduction of core granule protein synthesis, with the exception of MPO (Fig. 3E). Collectively, these data argue for a significant perturbation of granule protein synthesis in cultured neutrophils, which is supportive of our suggestion that they have not reached complete maturity.

### CRISPR/Cas9 genome editing of cultured neutrophils

Neutrophils are notoriously short lived and difficult to transfect, therefore the ability to culture them from CD34^+^ stem cells offers a window of opportunity to explore gene editing [44]. We investigated whether cultured neutrophil precursors are amenable to genetic manipulation, prior to differentiation, as a tool for modifying gene expression in immature neutrophils. We targeted CD34^+^ HSPCs (day 3 of culture) and used nucleofection to deliver ribonucleoproteins (RNPs) of Cas9 and guide RNA (gRNA). We used two different gRNAs, targeting β_2_ microglobulin (β_2_M), a transmembrane protein expressed on all nucleated cells, which is part of the major histocompatibility complex, and CD11b, an integrin expressed on myeloid cells. Nucleofection was efficient and did not require selection, with differentiated neutrophils demonstrating 92.5% and 88.1% loss of β2M (Fig. 4A) and CD11b (Fig. 4B) surface expression, respectively, at culture endpoint (day 17). We found no significant difference in viability of the gene edited cells (Fig. S3A). Moreover, CD66b expression and therefore neutrophil differentiation was not affected (Fig. S3B), confirming that modification of precursors by CRISPR/Cas9 is well tolerated and can be used as a molecular tool to investigate immature neutrophils.

**Fig. 4.**
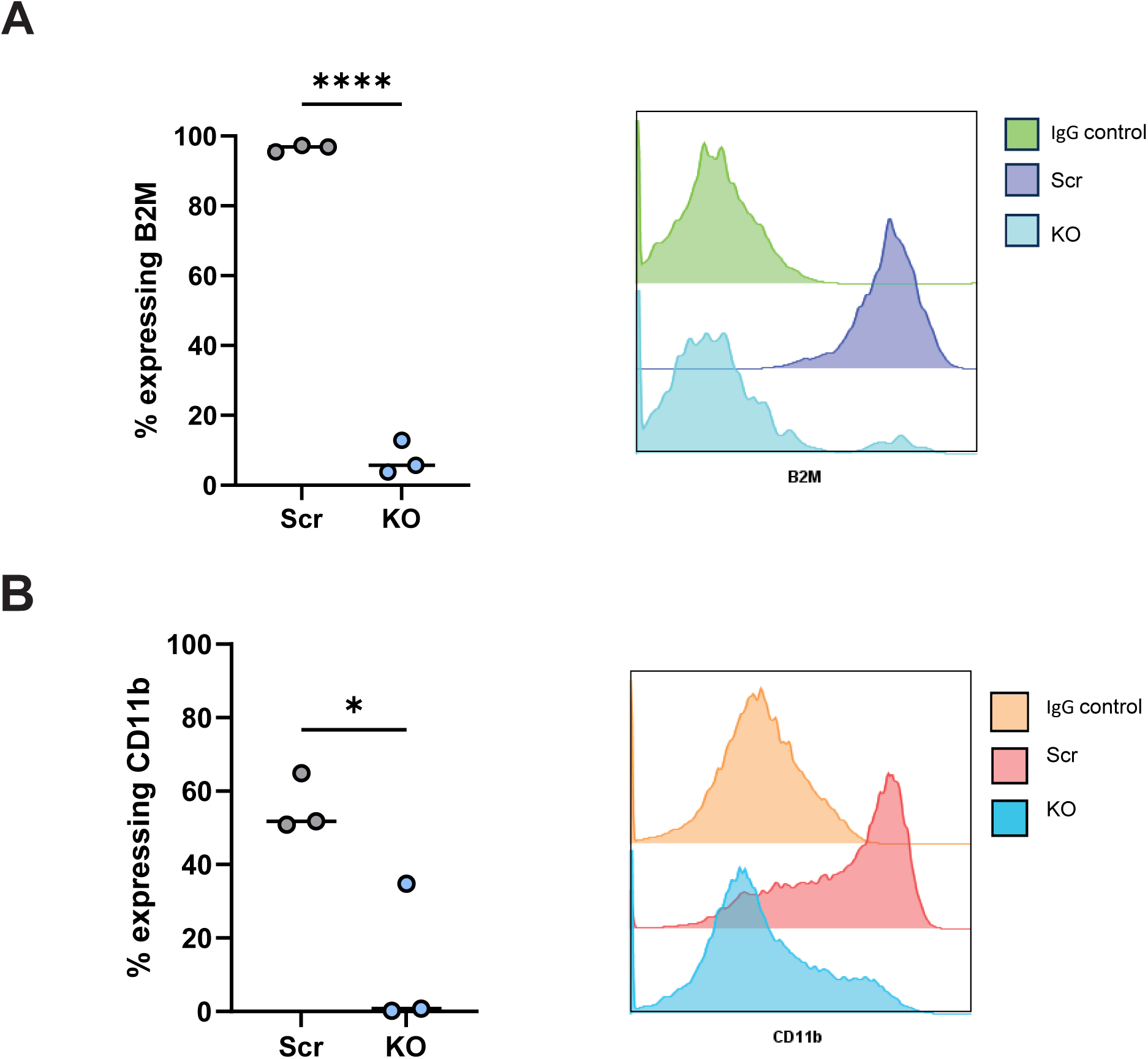
Genome editing of cultured neutrophils. CRISPR/Cas9 mediated deletion of A) *β_2_M* and B) *CD11b* demonstrated by flow cytometric quantification of the percentage of cells expressing targeted protein (left) and representative histograms of differentiated neutrophils (D17, right), n=3-4. Data were analysed by two tailed students t test, * = p<0.05, ** = p< 0.01, *** = p< 0.001, ****=p<0.0001.

## Discussion

Many basic questions in neutrophil biology remain unanswered, despite the importance of these cells in antimicrobial defence and inflammatory disease. This is in part because, neutrophil research is hampered by a lack of tools for genetic manipulation and by the short lifespan of neutrophils isolated from peripheral blood. One important outstanding question is the existence of neutrophil heterogeneity in disease conditions and whether neutrophils produced during inflammation differ from those produced at steady state. Studies using single cell RNA sequencing are providing compelling evidence for the presence of multiple neutrophil states [19]. Unsurprisingly for a cell with a short circulating lifespan (1-5 days in circulation) [1], neutrophil maturity is emerging as a major factor in determining phenotype and function.

Building on previous work, we developed an improved method for *ex vivo* culture and genetic manipulation of neutrophils. Our optimised protocol uses CD34^+^ stem cell cells isolated from waste apheresis cones, rather than the more restricted and costly embryonic or induced pluripotent stem cells, or HSPCs isolated from cord blood or bone marrow [31, 32, 37–40]. Compared to previous reports [30, 32, 34, 37, 40, 45–47], our protocol offers the combined advantages of 1) a shortened 17-day differentiation, 2) high levels of purity and 3) high yield. Most recently, Kuhikar et al. described neutrophil differentiation from apheresis cones, showing an average 72.4-fold expansion, with 57.37% of neutrophils expressing CD66b and 70.48% expressing CD15 [48]. Our protocol achieves an average 326-fold expansion with 75.45% of neutrophils co-expressing CD66b and CD15. The yields obtained using this culture method are robust, however there is significant variation between apheresis blood donors. We currently do not know the reason for this but assume it is natural biological variation in the ability of donor CD34^+^ stem cells to proliferate under our standardised conditions.

Quantification of expression of the well-established maturity markers CD10 and CD101 demonstrated that cultured neutrophils phenotypically resemble immature neutrophils mobilised by GCSF administration. Similarly to GCSF-D neutrophils [18], cultured neutrophils demonstrated reduced *C. albicans* killing and overproduction of IL-6, compared to steady state native cells. The overproduction of IL-6 may be a potential pathogenic mechanism in diseases where immature neutrophils are implicated. Indeed, secretion of IL-6 by immature neutrophils has been implicated in autoinflammatory diseases such as chronic graft versus host disease and adult-onset Still’s disease [49, 50].

Cultured neutrophils produce an efficient NOX2 oxidative burst, as previously shown [32, 33, 36, 40, 47, 51], and have comparable levels of MPO activity to native cells. Interestingly MPO was enriched in the proteome of cultured cells, reflecting reports of elevated MPO expression in immature CD10^-^ neutrophils circulating in myocardial infarction patients [52].

In contrast to MPO, many other granule proteins were significantly reduced in the proteome of cultured neutrophils. This paucity of cytoplasmic granule proteins was observed for primary (NE), secondary (lactoferrin) and tertiary (matrix metalloprotease 9 (MMP9), CRISP3) granules, and may explain the defect in *C. albicans* killing. Degranulation has been the least studied antimicrobial response in cultured neutrophils, with only Dick *et al.* reporting reduced primary granule exocytosis in cultured versus native neutrophils [39], in line with our finding of reduced NE release. It is likely that both findings are explained by perturbations in granule synthesis rather than reduced function of exocytosis machinery. The decrease in granule protein abundance and the maintenance of mitochondrial metabolism are both indicative of incomplete maturation of cultured neutrophils. This is reminiscent of *in vitro* erythroid culture from CD34^+^ HSPCs, where the majority of the differentiated cells are immature erythrocytes (reticulocytes), which subsequently mature in circulation [53, 54].

*Ex vivo* culture presents opportunities for genome editing that are not possible in neutrophils isolated from peripheral blood. Similar to previous knockout [44] and overexpression efforts [38], we show that CRISPR/Cas9 can be used to modulate gene expression in cultured neutrophils. Nucleofection of ribonucleoprotein negates the need for using plasmid constructs, which we have found can impact neutrophil differentiation. This represents an important advance in the manipulation of cultured neutrophils that will facilitate moving away from use of imperfect immortalised neutrophil-like cell lines and mouse models, which do not fully recapitulate human neutrophil phenotypes and functions [55]. This technique also provides a model system for investigating the molecular bases of neutropenia and neutrophil immunodeficiencies caused by germline mutations.

In summary, cultured neutrophils can be used to model immature, bone marrow-mobilised neutrophils. *Ex vivo* differentiation, coupled with genome editing, are useful new tools for increasing our understanding of the predominant state of neutrophil circulating in patients with systemic inflammatory disease.

## Materials and methods

### CD34^+^ stem cell isolation

Peripheral blood mononuclear cells (PBMCs) were isolated from apheresis waste products (NHSBT, Filton, Bristol, UK) with ethical approval from NHS Research Ethics committee (REC 18/EE/0265). PBMCs were isolated by density centrifugation using Histopaque p1077 (Sigma Aldrich) as previously described [56, 57]. 8-10mL of blood from apheresis were mixed with 60µL citrate-dextrose solution (ACD, Sigma Aldrich). 10mL of Hanks’ balanced salt solution (HBSS, Lonza) was added to samples and the mixture was layered over 25mL of room temperature (RT) Histopaque p1077 (Sigma Aldrich) then centrifuged at 400g at RT for 35 minutes with no brake. The interface layer that contains PBMCs was collected and washed five times with HBSS supplemented with ACD (0.6% v/v, Sigma Aldrich). Finally, cells were resuspended in 10mL red cell lysis buffer (55mM NH4Cl, 0.137mM EDTA, 1mM KHCO3, pH 7.5 in water) and washed once with HBSS supplemented with ACD (0.6% v/v, Sigma Aldrich). CD34^+^ cells were isolated from PBMCs using a Human CD34 MicroBead Kit (Miltenyi Biotec) and LS columns (Miltenyi Biotec) as outlined in the manufacturer’s instructions.

### Neutrophil differentiation

CD34^+^ cells were cultured in IMDM (Biochrom, Source Biosciences, Cambridge UK) supplemented with 10% (v/v) Fetal Calf Serum (FCS, Life Technologies) and 1% (v/v) Penicillin/Streptomycin (P/S, Sigma Aldrich), at 37°C in 5% CO_2_. Stem Cell Factor (SCF, 50ng/mL, Miltenyi Biotech), Flt-3 Ligand (50ng/mL, Miltenyi Biotech), Interleukin-3 (IL-3, 10ng/mL, R&D Systems), GM-CSF (10ng/mL, Miltenyi Biotech) G-CSF (10ng/mL, Miltenyi Biotech) were introduced at the following times post CD34^+^ isolation: IL-3, SCF and Flt3-L from day 0-3; GM-CSF, IL-3, SCF and Flt3-L from day 3-7; GM-CSF and G-CSF from day 7-10 and G-CSF only from day 10-17. CD34^+^ cells were initially plated at 0.1-0.2×10^6^ cells/mL on Day 0. A full media change was completed at Day 3 post CD34^+^ isolation, after which point cultures were supplemented with additional media every 2-3 days to maintain a cell density of 0.5×10^6^ cells/mL. Unless stated otherwise all functional assays were completed between Day 17 and Day 19 of culture.

### Native neutrophil isolation from peripheral blood

Blood samples from consented healthy donors at the University of Bristol were collected with ethical approval from NHS Research Ethics committee (REC 18/EE/0265), into EDTA tubes. Neutrophils were isolated using the EasySep^TM^ direct human neutrophil isolation kit (STEMCELL Technologies) as per the manufacturer’s instructions.

### Multi-fluorophore flow cytometry analysis

Flow cytometry analysis was conducted using 5×10^5^ Day 17-19 cultured or GCSF-D neutrophils as previously described [20]. Briefly, samples were washed with PBS, resuspended in 0.1% Zombie Aqua live/dead stain in PBS (BioLegend) and incubated for 10 minutes in the dark at RT. Samples were then incubated in FC Block (BioLegend) diluted in flow buffer (5mM ETDA and 0.5% bovine serum albumin (BSA) in PBS) on ice for 5 minutes, after which a master mix of primary antibodies was added and incubated for 30 minutes on ice protected from light. Samples were washed twice with flow buffer and fixed using 2-4% paraformaldehyde for 20 minutes at RT. Cells were analysed using a BD X20 Fortessa flow cytometer within 7 days of sample preparation. Appropriate single fluorescence colour compensation controls were conducted in parallel using Invitrogen OneComp eBeads (Thermo Fisher Scientific). At least 10,000 events recorded per sample, gated on a singlet, live population and data were processed in FlowJo software (version 9).

### C. *albicans* killing assay

C. *albicans* CaSS1 strain was grown overnight in Yeast Extract-Peptone-Dextrose (YPD) medium supplemented with 0.05 µg/ml doxycycline at 30°C, 200 RPM in 5% CO_2_ for 16 hours. The next day, C. *albican*s concentration was determined by optical density, sub-cultured in YPD with 0.05 µg/ml doxycycline for 3 hours at 30°C, 200 RPM in 5% CO_2_. Fungi were resuspended in RPMI-1640 and opsonised with 5% pooled human serum for 30 minutes before incubating with native or cultured neutrophils at a MOI of 2.5 or 5 at 37°C for 2.5 hours. Samples were treated with 0.1% Triton X-100 (Sigma Aldrich) to lyse neutrophils, washed three times with PBS, incubated with alamarBlueTM (Thermo Fisher Scientific) for 17 hours and fluorescence was measured using a FLUOstar Omega Microplate Reader (BMG Labtech) to quantify metabolically active *C. albicans*.

### Cytokine release

1×10^5^ neutrophils were plated in 200µl in triplicate in RPMI-1640 with phenol red (GIBCO), 10% fetal bovine serum (FBS, Sigma) and 1% P/S (Biochrom) and stimulated with 100ng/mL bacterial lipopolysaccharide (LPS from *E. coli* O127:B8, Sigma Aldrich) or 5 µM resiquimod (Sigma Aldrich) overnight at 37° C in 5% CO_2_. IL-8 and IL-6 levels in the resulting supernatants were measured using Human IL8/CXCL8 DuoSet ELISA and Human IL-6 DuoSet kits following the manufacturers protocol (both R&D Systems).

### Zymosan Degranulation Assay

1×10^5^ neutrophils were plated in duplicate, in 200µl RPMI-1640 (GIBCO) supplemented with 0.025% human serum albumin (HSA; Sigma-Aldrich) and 10mM HEPES (Sigma-Aldrich) and were stimulated with 25µg/mL opsonised Zymosan (Sigma Aldrich). After one hour, samples were centrifuged 300g, 100µL of supernatant removed from each well and the NE concentration measured using a Human Neutrophil Elastase/ELA2 DuoSet ELISA kit (R&D Systems) as per the manufacturer’s protocol.

### APF Reactive oxygen species production

1×10^5^ neutrophils in 100ul of ROS media (HBSS (Lonza) with 10mM HEPES and 0.025% HSA, both Sigma Aldrich) supplemented with 10µM APF (Thermo Fisher) in black, clear, flat-bottomed plates. Cells were incubated at 37°C, 5% CO_2_ for 45 minutes, centrifuged at 400g for 5 minutes, media aspirated and replaced with plain ROS media. Neutrophils were stimulated with 100nM PMA (Sigma Aldrich), and fluorescence measured every 2.5 minutes for 4 hours using a BMG FLUOstar plate reader (emission: 490nm, excitation: 515nm).

### TMT Mass spectrometry

Day 18 cultured neutrophils and native neutrophils were isolated and prepared from 3 separate donors as described above. Sample pairs were donor-matched, with CD34^+^ cells and native neutrophils isolated from the same fresh apheresis cone. Neutrophils for analysis were sorted against CD66b expression using a BD Influx Cell Sorter (BD Biosciences). Cells were immediately pelleted and lysed in supplemented RIPA buffer (EDTA 10 mM, 50 mmol/l TCEP (Sigma Aldrich), 2mM PMSF protease inhibitor, 1/50 v/v Protease Inhibitor Cocktail Set V (Calbiochem)) and flash-frozen in liquid nitrogen for storage until later analysis. All samples were then thawed on ice, sonicated to fragment DNA, and measured for protein concentration with a Pierce® BCA Protein Assay Kit (Thermo Scientific, cat no: 23227) according to the manufacturer’s instructions. Due to TCEP being used during cell lysis, a reducing agent-compatible kit was used. 100 µg of each sample were then digested with trypsin and labelled with Tandem Mass Tag (TMT) reagents according to the manufacturer’s protocol (Thermo Fisher Scientific). The resulting peptides were identified by nano LCMS/MS with a Orbitrap Fusion Tribrid Mass Spectrometer (Thermo Fisher Scientific). Raw files were analysed using Proteome Discoverer software v. 2 and cross-referenced against the human UniProt database (human). PD analysis was conducted for dull trypsin digestion, removing all hits with more than one missed cleavage. All peptides were filtered to meet a false discovery rate (FDR) of 1%. Log_2_ fold changes (Log2FC) were calculated between cultured and native neutrophils to identify differentially expressed proteins (|Log_2_FC| > 1, P value < 0.05). Log_2_FC volcano plots were generated using Microsoft Excel. Log_2_FC clustering analysis and the resulting heatmap visualisations were generated using R v. 4.2.2. Protein-protein interaction networks for differentially expressed proteins were generated using the STRING database and Cytoscape v. 3.9.1.

### CRISPR/Cas9 gene editing

Nucleofection of ribonucleoproteins (RNP) was completed using the Nucleofector 4D (Lonza) using a P3 Primary Cell 4D-NucleofectorTM X Kit S (Lonza) and Invitrogen™ TrueCut™ Cas9 Protein v2 (Thermo Fisher Scientific) following the manufacturer’s recommended protocols. CD34^+^ cells were isolated and cultured as described above until Day 3 of culture. 0.3×10^6^ cells were transduced per reaction. 50 pmol of Cas9 was mixed with 125pmol of gRNA (62.5pmol of two guides with the same gene target or a scrambled (SCR) gRNA control) per reaction and incubated at 25°C for 15 minutes to form RNPs, which were stored on ice for up to 4 hours. All guides were designed using the Synthego CRISPR Design Tool and produced by Synthego (California, USA). Supplemented Nucleofector Solution (SNS) was made up fresh at a 4.5:1 ratio of nucleofector solution to supplement. 0.3×10^6^ Day 3 cells were spun down, washed in PBS, and resuspended in 20µL SNS. Cells were added to RNPs, mixed gently, and transferred to a nucleocuvette cassette. Cells were then electroporated using the manufacturers recommended Nucleofector 4D program EO-100. 80µL of prewarmed 37°C StemSpan was dripped gently into each reaction to dilute the SNS. Cells were replated in 2mL of StemSpan (Stem Cell Technologies) supplemented with 1% (v/v) P/S (Sigma Aldrich), SCF (50ng/mL, Miltenyi Biotech), Flt3-L (50ng/mL, Miltenyi Biotech), IL-3 (10ng/mL, R&D Systems), GM-CSF (10ng/mL, Miltenyi Biotech) and GCSF (10ng/mL, Miltenyi Biotech). Cells were allowed to recover for 48 hours, after which they were cultured from Day 5 as indicated in the neutrophil culture methodology section (Fig. S1).

### Statistical analysis

Data was organised and analysed using GraphPad Prism 8 software. Mean ± SD is plotted for a minimum of n=3 differentiations unless otherwise stated. Statistical analysis was completed where appropriate using student’s t test when comparing two different samples, or one-way ANOVA where multiple samples were being examined, with asterisks on the graph represent the following: \**P* < 0.05, \*\**P* < 0.01, and \*\*\**P* < 0.001.

## Author contributions

**MT** and **KR** optimised the CD34^+^ neutrophil culture system and performed proteomics experiments. **CN, KR, CR,** and **NP** performed experiments and analysed data; **PM** analysed proteomics data and generated figures; **KF** organised human sample collection; **SD** supervised fungal experiments; **BA and AT** conceived and supervised the study. **CN** and **BA** wrote the manuscript.

All authors read, provided input and approved the final manuscript.

## Funding statement

This work in BA’s lab was funded MRC grant MR/R02149X/1. AT was funded by a NHS Blood and Transplant (NHSBT) R&D grant (WP15-05) and a National Institute for Health Research Blood and Transplant Research Unit (NIHR BTRU) in Red Blood Cell Products at the University of Bristol in partnership with NHSBT (IS-BTU-1214-10032) (AT). KR is funded by a Wellcome Trust Dynamic Cell PhD studentship. The views expressed are those of the authors and not necessarily of the NHS, the NIHR or the Department of Health.

## Data Availability

Data are freely available upon request to the corresponding authors.

## Acknowledgments

We thank all the blood donors for participating in our study. We acknowledge Przemek Zakrzewski and Sarah Groves for technical assistance and James Griffin (NHSBT, Filton, Bristol) for clinical samples.

## Supplemental Figures

**Fig. S1.**
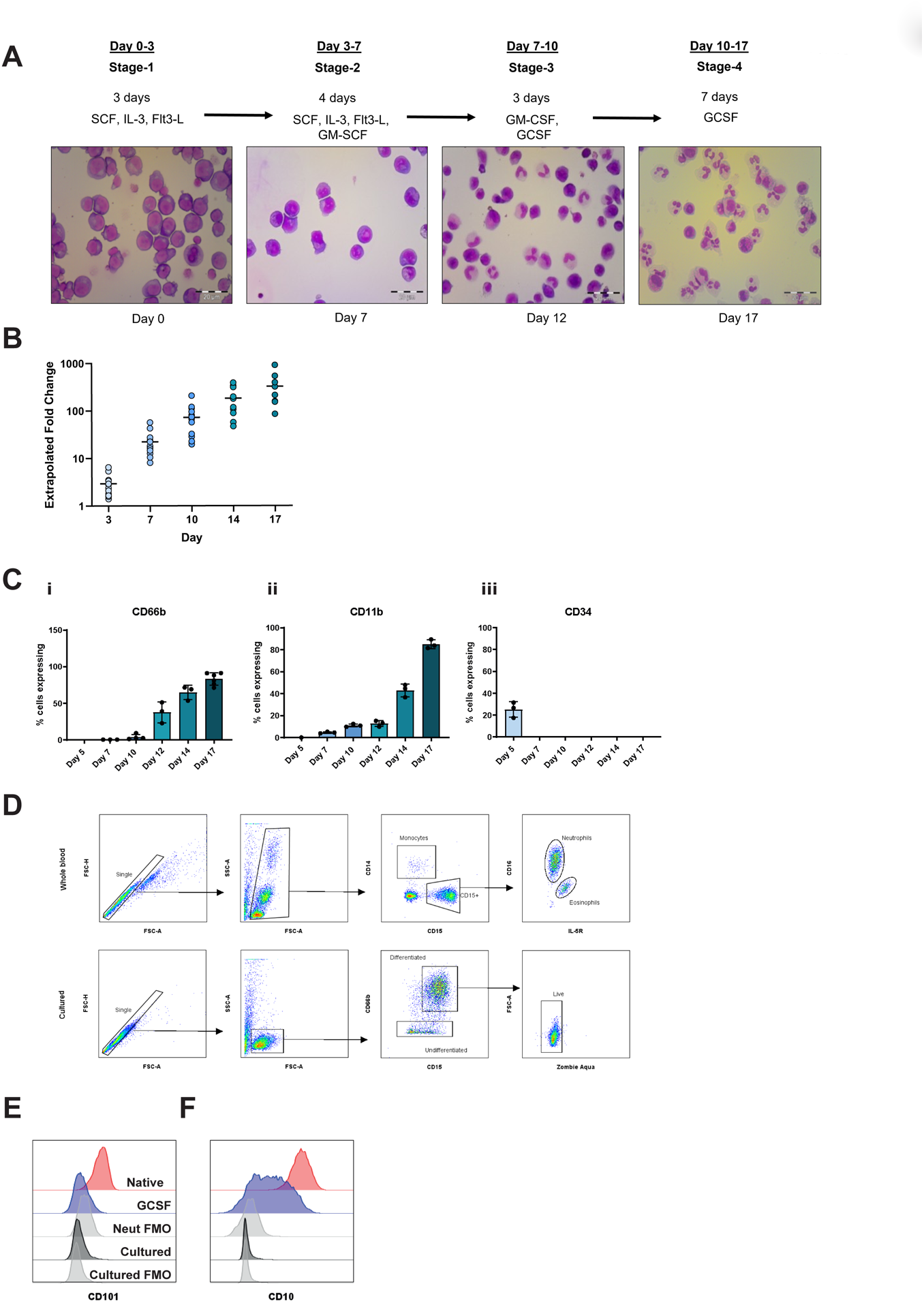
Optimisation of neutrophil culture protocol. A) Representative Wright Giemsa-stained cytospin images of neutrophil differentiation steps with cytokine protocol. B) Extrapolated fold expansion of cultured neutrophils over 17 days of differentiation, n=7-11. C) Surface marker expression of HSPC marker CD34 (i) and granulocyte markers CD66b (ii) and CD11b (iii) over 17 days of differentiation, n=3. D) Representative gating strategy of singlet, appropriately sized neutrophil populations. E) CD101 and F) CD10 expression in peripheral blood neutrophils from native (red), GCSF-D (blue) and cultured neutrophils (black). Fluorescence minus one (FMO) controls for cultured neutrophils and peripheral blood neutrophils are displayed (grey).

**Fig. S2.**
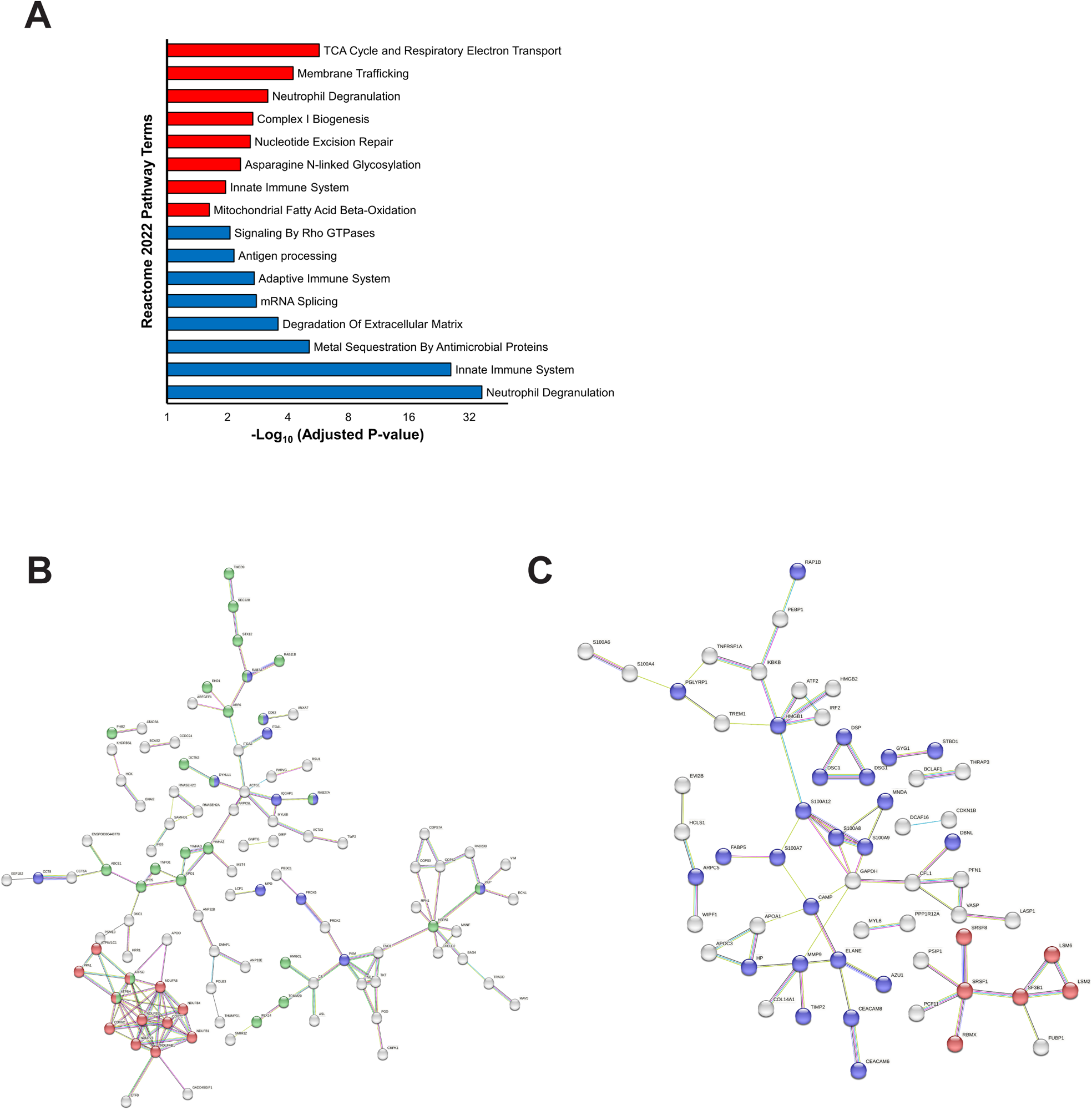
STRING analysis of upregulated and downregulated proteins in cultured compared to native neutrophils. A) Top 8 unique Reactome 2022 pathway terms of enriched (red) and under-represented (blue) genes using the GO Biological Pathway module. Enriched (B) and under-represented (C) proteins processed using the STRING database using a medium confidence setting to produce protein-protein interaction networks. Clusters are shown in different colours: A) ribosomal proteins in red, mitochondrial in blue and degranulation proteins in green and in B) innate immune system and degranulation in red, mRNA processing in blue and chromatin organisation in green.

**Fig. S3.**
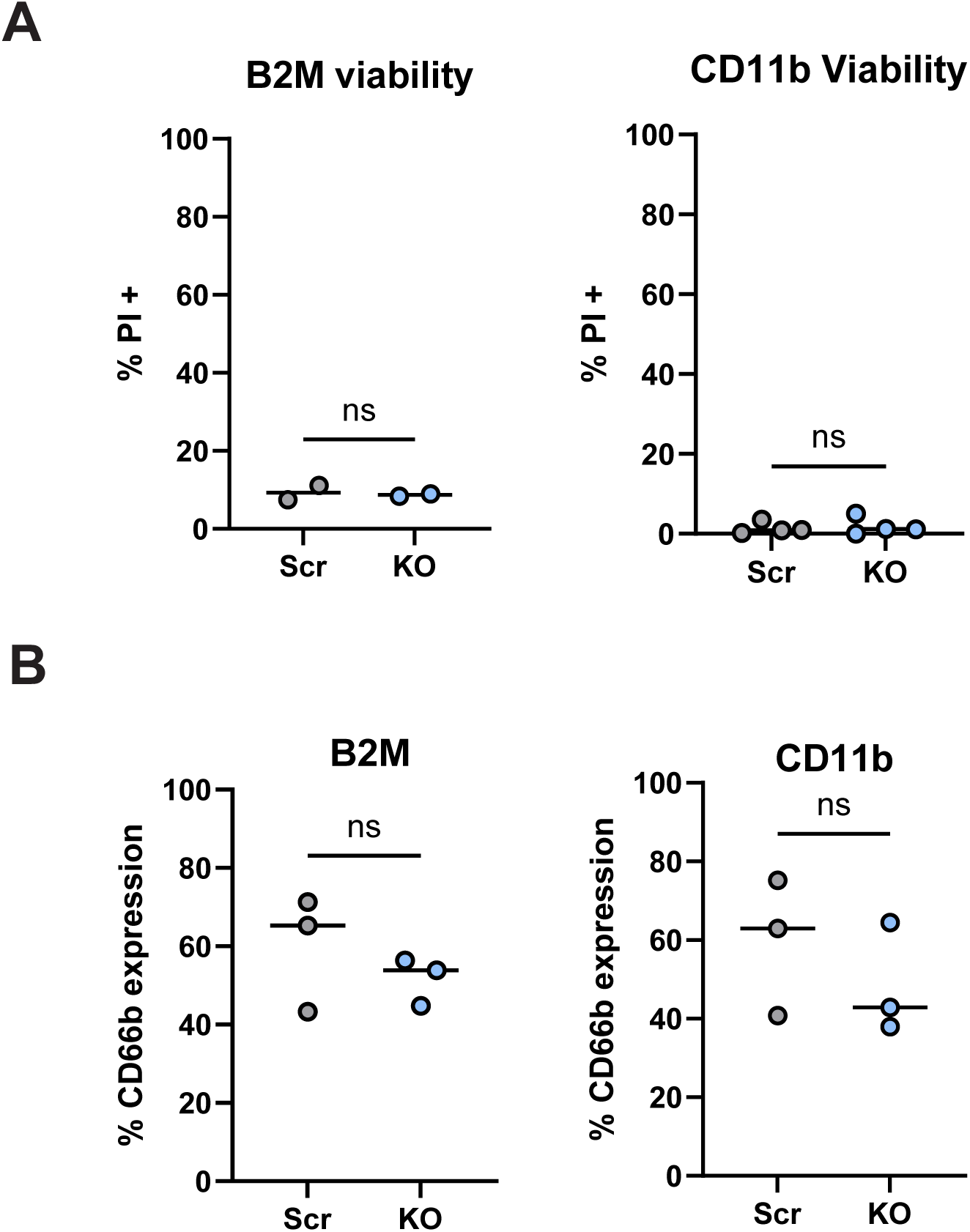
CD66b expression and viability is unchanged by CRISPR/Cas9 mediated knockout in cultured neutrophils. A) Percentage of propidium iodide positive cells in Scr vs *β_2_M* (left) and *CD11b* (right) KO cells respectively by flow cytometric surface marker staining on day 7 of culture. B) The percentage of CD66b expressing cells in *β_2_M* (left) and *CD11b* (right) CRISPR/Cas9 KO cells, by flow cytometric surface marker staining on day 17 of culture.

## Supplemental Results

**Supplementary Table 1:** Proteins uniquely expressed in cultured neutrophils and not native neutrophils.

| Gene name | Gene description |
| --- | --- |
| ANXA6 | Annexin A6 |
| DNAJC7 | DnaJ homolog subfamily C member 7 |
| EIF5 | Eukaryotic translation initiation factor 5 |
| DKFZp686E1893 | Putative uncharacterized protein DKFZp686E1893 |
| CCNB2 | G2/mitotic-specific cyclin-B2 |
| SELENOT | Selenoprotein T |
| ARL2 | ADP-ribosylation factor-like protein 2 |
| SPCS3 | Signal peptidase complex subunit 3 |
| PPDPF | Pancreatic progenitor cell differentiation and proliferation factor |
| MT1G | Metallothionein-1G |
| PNKD | Probable hydrolase PNKD |
| KCNK1 | Potassium channel subfamily K member 1 |

## Supplemental Methods

### Cytospin preparation and imaging

1×10^5^ cells were removed from cultures at the indicated timepoints post CD34^+^ isolation. Cells were washed with PBS and spun onto glass slides at 1000g for 5 minutes (Thermo Scientific Cytospin). Samples were then fixed in 100% methanol for 10 minutes and then stained with May Gruwald–Giesma stains as per the manufacturer’s instructions (Merck).

### Four fluorophore flow cytometry analysis

1×10^5^ cells were removed on day 8 of cell culture and labelled with conjugated antibodies for 25 minutes at 4°C, using commercial nonspecific IgG controls for nonspecific staining. Propidium iodide labelling was used to identify the dead cell population. Samples were analysed using a MacsQuant flow cytometer (Miltenyi Biotec) and processed using FlowJo software (Version 9).

## Supplemental Tables

**Supplementary Table 2:**
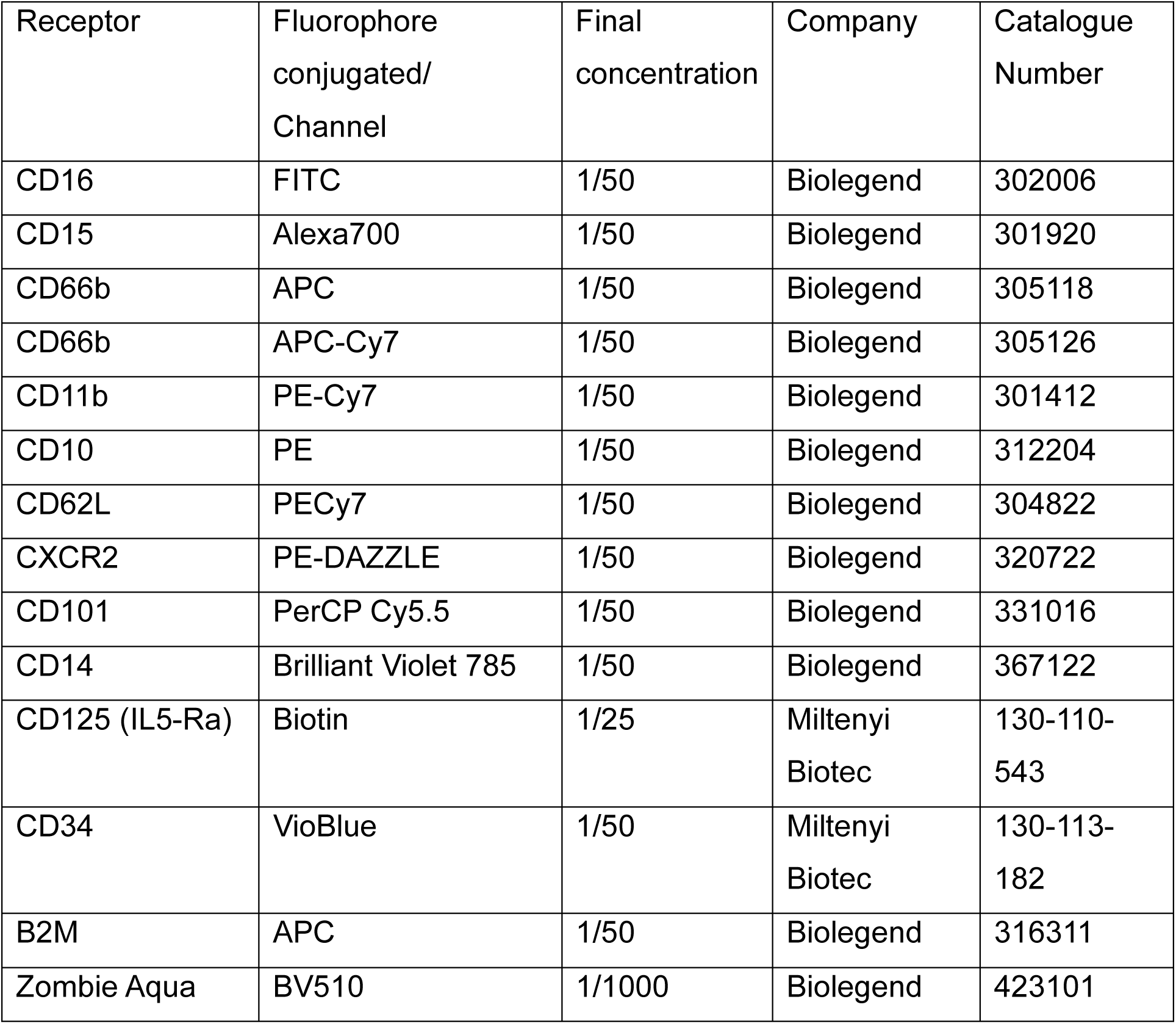
Antibodies and fluorescent dyes used for flow cytometry.

**Supplementary Table 3:**
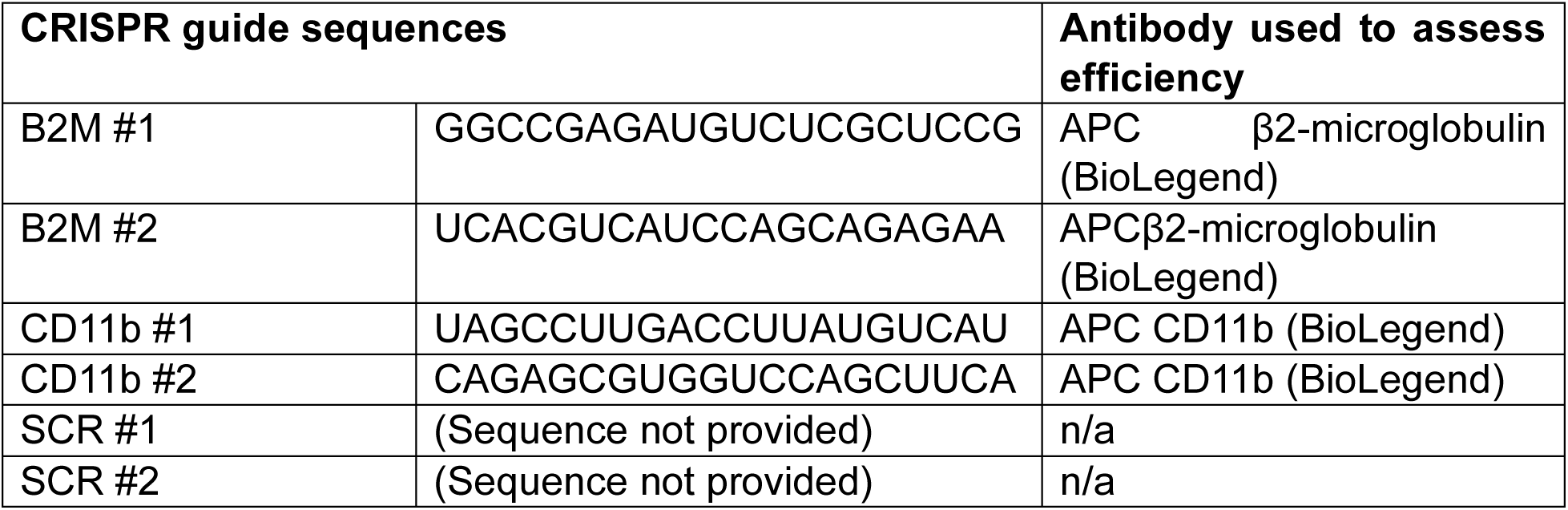
Synthego guide RNA for CRISPR-Cas9 gene editing.

## Notes

### Competing Interest Statement

The authors have declared no competing interest.

### Funding Statement

This work in the lab of BA was funded MRC grant MR/R02149X/1. AT was funded by a NHS Blood and Transplant (NHSBT) R&D grant (WP15-05) and a National Institute for Health Research Blood and Transplant Research Unit (NIHR BTRU) in Red Blood Cell Products at the University of Bristol in partnership with NHSBT (IS-BTU-1214-10032) (AT). KR was funded by a Wellcome Trust Dynamic Cell PhD studentship. The views expressed are those of the authors and not necessarily of the NHS, the NIHR or the Department of Health.

### Author Declarations

The NHS Research Ethics Committee gave ethical approval for this work (REC 18/EE/0265).

